# Single-cell Profiling of the Response to Poly I:C in Peripheral Blood Mononuclear Cells in Severe Asthma

**DOI:** 10.1101/2020.09.21.20197863

**Authors:** Ailu Chen, Maria P. Diaz-Soto, Miguel F. Sanmamed, Taylor Adams, Jonas C. Schupp, Amolika Gupta, Clemente Britto, Maor Sauler, Xiting Yan, Qing Liu, Gustavo Nino, Charles S. Dela Cruz, Geoffrey L. Chupp, Jose L. Gomez

## Abstract

**Background:** Asthma has been associated with impaired interferon responses. Multiple cell types have been implicated in these impaired responses and may be responsible for increased exacerbations and immunopathology of asthma.

**Objective:** Characterize the single-cell response to Poly I:C of peripheral blood mononuclear cells (PBMCs) of patients with severe asthma (SA).

**Methods:** Two complementary single-cell methods, DropSeq for single-cell RNA sequencing (scRNA-Seq) and mass cytometry (CyTOF), were used to profile PBMCs of SA and healthy controls (HC). Poly I:C and unstimulated cells were analyzed in this study.

**Results:** PBMCs (n=9,414) from five SA (n=6,099) and three HC (n=3,315) were profiled using scRNA-Seq. Six main cell subsets, including CD4+ T cells, CD8+ T cells, natural killer (NK) cells, B cells, dendritic cells (DCs), and monocytes, were identified. CD4+ T cells were the main cell type and demonstrated a pro-inflammatory profile characterized by increased *JAK1* expression in unstimulated cells. Following Poly I:C stimulation, PBMCs from SA had a robust induction of interferon pathways compared with HC. Additional analyses to identify core regulators of the enhanced interferon response in SA identified *IRF1, STAT1, IRF7, STAT2*, and *IRF9*. CyTOF profiling of Poly I:C and unstimulated PBMCs (n=120,000) from the same individuals (SA=4; HC=2) demonstrated higher numbers of CD8+ effector cells and Th1 CD4+ T cells in unstimulated conditions, followed by a decrease of these two cell subsets after poly I:C stimulation.

**Conclusion:** Single-cell profiling of PBMCs with scRNA-seq and CyTOF in patients with SA identified activation of pro-inflammatory pathways at baseline and strong response to Poly I:C, as well as quantitative changes in CD8+ effector cells and Th1 cells. Thus, transcriptomic and cell quantitative changes are associated with immune cell heterogeneity in severe asthma.

**Key Messages:**

- Single-cell RNA sequencing identified a pro-inflammatory status in unstimulated PBMCs of severe asthmatics.
- Mass cytometry identified quantitative differences in CD8+ effector cells and Th1 cells of severe asthmatics.
- The response to Poly I:C stimulation, an interferon agonist, was not impaired in a subgroup of patients with severe asthma.

**Capsule summary:** Single-cell profiling of PBMCs in severe asthmatics characterized gene expression responses to an interferon agonist and quantitative differences in distinct cell populations. Comprehensive single-cell immune may help identify key cell features responsible for asthma heterogeneity.

## Introduction

Asthma endotypes, disease subtypes defined by a molecular mechanism or a treatment response, have been associated with cell abundance, including eosinophils ^1–3^. However, the cellular heterogeneity in immune cells within and between patients with asthma is not only restricted to absolute cell counts ^3^, but also includes distinct transcriptional changes in the airway ^4–6^ and blood ^7–9^. The collective transcriptomic signatures derived from bulk genome-wide RNA analyses support dysregulated gene expression in disease pathogenesis.

Despite these advances, a significant limitation of bulk RNA analyses is that averaging transcriptomes can result in the loss of cell-specific signals that may underlie disease endotypes. Furthermore, innate and adaptive immune cellular responses are involved in asthma pathogenesis, and specific disease phenotypes result from the interaction of immune cells with the environment ^10^. Among environmental influences, increased susceptibility to viral infections is a feature strongly associated with asthma, with several studies showing impairment in different components of the antiviral response ^9,11–17^. Although these studies have informed our understanding of asthma pathogenesis, we lack a comprehensive picture of functional and quantitative responses at the single-cell level in severe asthma (SA), limiting our understanding of the contribution of distinct cell subtypes to this disease phenotype.

Based on these observations, we hypothesized that specific peripheral blood mononuclear cells (PBMCs) of SA have an impaired response to interferon (IFN). We used two complementary single-cell methods to study transcriptomic and quantitative changes in PBMCs before and after stimulation with polyinosine–polycytidylic acid (poly I:C). Poly I:C, a synthetic double-stranded RNA (dsRNA) analog and TLR3 agonist, leads to the production of type I interferons (IFNs) ^18^, and compared these profiles with unstimulated PBMCs. We used DropSeq for single-cell RNA sequencing (scRNA-seq) ^19^ and mass cytometry (CyTOF) ^20^ to profile single cells in PBMCs of patients with SA and healthy controls (HC), to determine whether cell-specific signatures are associated with a decreased response to IFN.

## Methods

### Sample Collection

Human donors were recruited as part of the GenEx study at the Yale Center for Asthma and Airway Disease (YCAAD). Whole blood was collected from 8 subjects (SA=5; HC=3) into BD Vacutainer® Sodium Heparin Tubes (BD Biosciences, Franklin Lakes, NJ). PBMCs were isolated using Lymphoprep and SepMate™-15(IVD) (STEMCELL Technologies, Vancouver, Canada) according to the manufacturer’s protocol. Isolated PBMCs were aliquoted and stored in cryotube vials (Thermo Scientific, Waltham, MA) with freezing medium (80% FBS, 20% DMSO) at a concentration of 5 × 10^6^ cells/mL. Cells were stored in Nalgene™ Cryo 1°C Freezing Container (Thermo Fisher, Rochester, NY) at -80°C overnight, then transferred into liquid nitrogen for long-term storage.

### Poly I:C Preparation

Poly I:C (InvivoGen, San Diego, CA) was reconstituted to 1 mg/mL stock solution per manufacturer’s protocol. Briefly, 10 mL of endotoxin-free physiological water was added into 10 mg poly I:C. The mixture was heated up to 70°C for 10 minutes, then was cooled down at room temperature for 1 hour. Poly I:C stock solutions were aliquoted into 100 µL each tube and stored at -20°C. 500 mL cell culture medium, which was composed of RPMI 1640 media (Gibco Laboratories, Gaithersburg, MD) supplemented with 10% heat-inactivated fetal bovine serum (MilliporeSigma, Burlington, MA) and 5% Penicillin-Streptomycin (MilliporeSigma, Burlington, MA), was prepared and aliquoted before the experiment started to ensure all the following experiments were using the same medium. A working solution at 10µg/mL poly I:C was prepared using cell culture medium 20 minutes before each use.

### *In Vitro* Exposure

One vial of frozen PBMCs (approximately 3 million cells) was removed from liquid nitrogen and quickly thawed in a 37°C water bath. Cells were then transferred into 10 mL pre-warmed cell culture medium and centrifuged at 400 g for 10 minutes to wash away DMSO. The supernatant was discarded after centrifugation. The cell pellet was resuspended with 3 mL 1x BD Pharm Lyse™ (BD Biosciences, Franklin Lakes, NJ) to remove red blood cells, incubated at room temperature for 3 minutes, then quenched with 7 mL PBS. An aliquot of cells was removed for cell counting before centrifuging at 400 g for 10 minutes. After centrifugation was completed, the supernatant was removed. Cells were resuspended with medium pre-warmed to achieve a concentration of 2 × 10^6^ cells/mL.

Cells were then plated into 96-well plates at 100 µL/well, and incubated at 37°C, 5% CO_2_ 100% humidity for 30 min. After incubation, cells were quenched with either 100 µL cell culture medium or 10 µg/mL poly I:C working solutions. The final concentrations were plain medium and 5 µg/mL poly I:C. Cells were cultured at 37°C, 5% CO_2_ 100% humidity for 24 hours.

### Drop-Seq Experiment

After 24 hours of incubation, cells were harvested into Eppendorf tubes and centrifuged for 10 min at 400g. Cells were washed once with 1 mL PBS with 0.01% BSA (PBS-BSA) to remove remaining FBS, then centrifuged for 8 min at 400g. After discarding the supernatant, cells were then resuspended with 1 mL plain PBS. An aliquot of cells was removed for cell counting and viability recording. Cells were adjusted to 140-160 live cells/µL using PBS-BSA as Drop-Seq input. The Drop-Seq experiment was carried out with the version 3.1 protocol from the McCarroll Lab ^19^. Briefly, in the microfluidic channel, the barcoded beads (Chem Genes, Wilmington, MA) encounter cells, and the oil flow separates each bead-cell into droplets. Once the cells were lysed within the droplets, released mRNAs were captured by bead barcoded sequences. In practice, beads (at 120 beads/µL), cells, and oil were pumped into a microfluidic system at speeds of 3,500 µL/hr, 3,500 µl/hr and 13,125 µL/hr respectively. Outflow droplets were collected in 50 mL Falcon tubes. After removal of excess oil from the bottom, outflow droplets were broken by adding 30 mL 6x Saline-Sodium citrate buffer (SSC) buffer (MilliporeSigma, Burlington, MA) and 1 mL of 1H,1H,2H,2H-Perfluoro-1-octanol (PFO) (MilliporeSigma, Burlington, MA) followed by forceful handshakes. Ambient RNA and excess oil were washed away by an additional 6x SSC buffer. Reverse transcription was conducted in the Eppendorf tubes. After reverse transcription, beads were washed with TE-SDS (10 mM Tris pH 8.0 +1 mM EDTA + 0.5% SDS) buffer and stored in TE-TW (10 mM Tris pH 8.0 +1 mM EDTA + 0.01% Tween-20) buffer at 4 °C before further steps.

Exonuclease treatment and PCR amplification were conducted within one week of reverse transcription of each sample. Beads were washed with water and then counted for PCR amplification. Apportion of 2,000 beads were put into one reaction; this would yield roughly 100 STAMPs. Multiple PCR batches from one sample were pooled before purification to ensure that at least 600 cells per sample were included. Purified PCR products were quantified using a high sensitivity bioanalyzer (Agilent 2100 expert High Sensitivity DNA Assay, Santa Clara, CA). cDNA library was constructed using 600 pg of each sample followed Drop-Seq customized tagmentation steps. Libraries were purified using the Agencourt AMPure XP system (Beckman Coulter, Brea, CA) before sequencing. RNA sequencing was conducted using Illumina Hi-Seq 2000 instrument in pair-end 75 bp. Control and poly I:C samples for each subject were pooled into one lane for sequencing at the Yale Center for Genome Analysis.

### CyTOF

One vial of frozen PBMC from human donors was cultured with poly I:C and cell culture medium as described above under *in vitro* exposure. Cultured cells were collected and transferred to 10 mL of pre-warmed DMEM (Gibco Laboratories, Gaithersburg, MD). Cells were centrifuged at 1,700 rpm for 7 minutes, resuspended in 1.5 mL of cell staining buffer (CSB) (Fluidigm, South San Francisco, CA), and 1-3 million cells were transferred to an Eppendorf tube for subsequent staining. Cells were then centrifuged at 1,700 rpm for 7 minutes, resuspended in 45 μL of CSB + 5 μL of Fc blocker (Biolegend, San Diego, CA) and incubated at room temperature for 10 min. Afterward, 50 μL of freshly prepared antibody cocktail was added (total staining volume 100 μL) and incubated on ice for 30 minutes. A summary of the antibodies used can be found in supplementary table 1. Next, cells were washed with CSB and stained for cell viability with 7.5 μM Cisplatin (Fluidigm, South San Francisco, CA) in RPMI 1,640 media for 1 minute and quenched with pure fetal bovine serum (MilliporeSigma, Burlington, MA). Cells were then washed with CSB, fixed, and permeabilized with the eBioscience Foxp3 Transcription Factor Staining Buffer Set (3:1 dilution) (Thermofisher Scientific, Waltham, MA) and stained with DNA intercalator (125 nM Iridium-191/193) overnight at 4°C. The next day, cells were washed first with CBS and then with ddH2O water (centrifuge speed 2100rpm for 10 min). Cells were then diluted to a concentration of 1×10^6^ cells/mL with a 1:7 dilution of beads solution (Fluidigm, South San Francisco, CA). Lastly, cells were filtered into a 35 μm cell strainer cap tube and were acquired at a rate of 300-500 cells/second using a CyToF2 mass cytometer (Fluidigm, South San Francisco, CA) at the Yale CyTOF facility. Figure 1 summarizes the study’s workflow.

**Figure 1.**
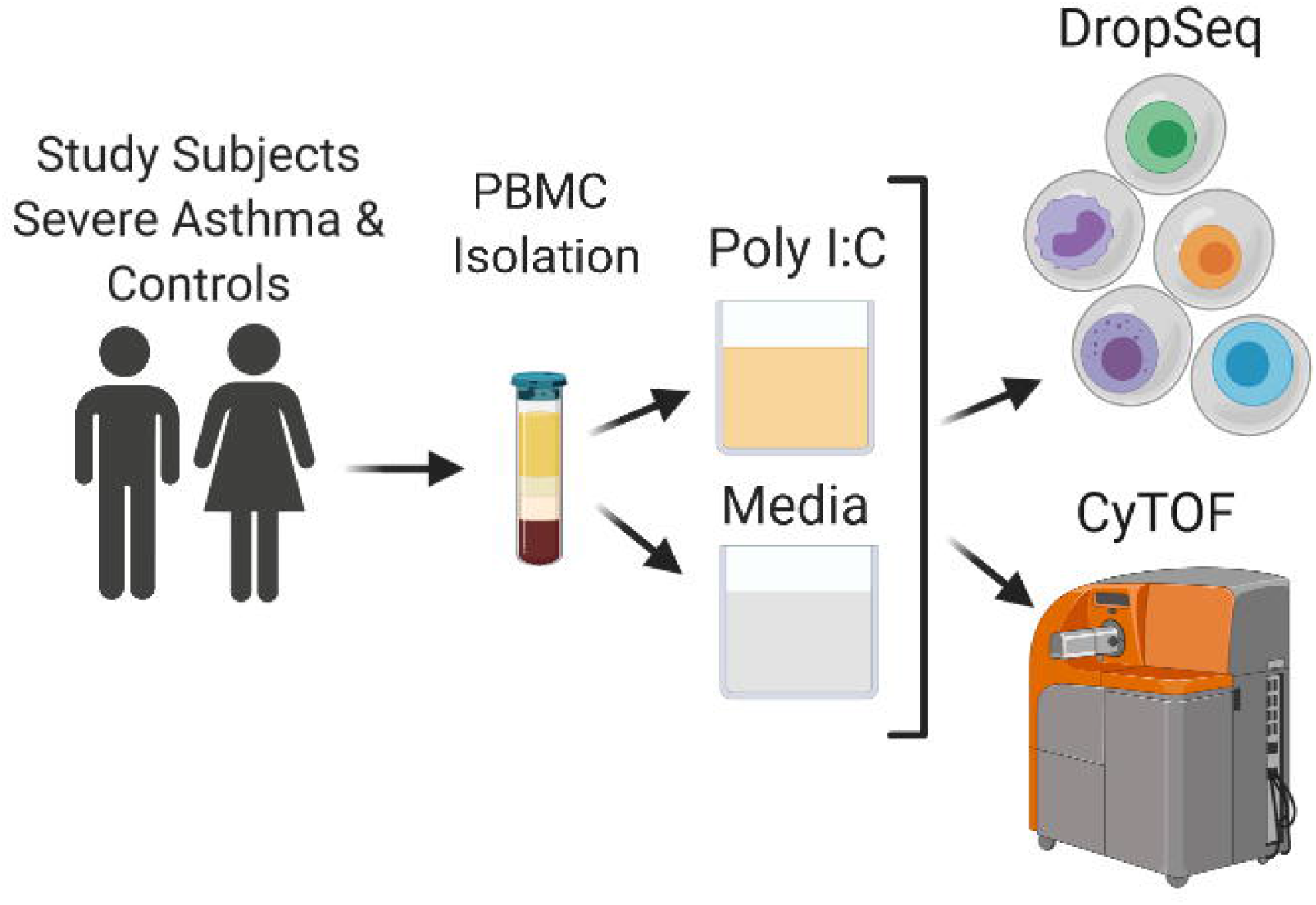
Study workflow.

### Data Analysis

Dropseq-derived sequencing reads were aligned to Genome Reference Consortium Human Build 38 (GRCh38) and then binned onto the cell barcodes corresponding to individual beads using Drop-Seq tools ^21^. We filtered out cells containing less than 200 genes and genes that were not detected in at least three cells to ensure qualifying genes and cells. Library-size normalization was performed with a global-scaling method by Seurat v 3.1.1 ^22^; briefly, the UMI-collapsed gene expression values for each cell barcode were scaled by the total number of transcripts and multiplied by 10,000. Data was then natural log-transformed before any further downstream analysis.

Highly variable genes were calculated and detected based on average expression and dispersion for each gene. Principal components analysis (PCA) was performed based on these highly variable genes. Statistically, significant PCs were selected for clustering analysis. The Uniform Manifold Approximation and Projection (UMAP) dimensional reduction technique was used to visualize the dataset. The positive and negative markers for each cluster were found through differential expression analysis, and only genes with expression percentage above 20% were kept. These markers were used for identifying cell identities. Differential gene expression was determined using the likelihood-ratio test for single-cell feature expression ^23^, using a fold change threshold of 10%. To identify changes between stimulated and unstimulated cells, we implemented the integration method described by Stuart and Butler et al. ^24^. P values were adjusted using Bonferroni correction, and only p<0.05 were reported and used in downstream analyses.

PHATE ^25^ embedding of CD4+ T cells was used to generate a pseudotemporal reconstruction of branching lineages^26^. Pseudotime reconstruction was followed by SCENIC analysis ^27^ to identify single-cell regulons. Briefly, a regulon is a set of genes in which a specific regulatory gene controls their expression. Predicted regulon activities per cell were calculated using the pySCENIC package with default settings. To this end, the cisTarget databases and the transcription factor motif annotation were used ^28^. The list of human transcription factors was obtained from the Aerts lab website ^29^.

Pathway maps, gene ontology (GO) processes, process networks, enrichment by diseases, and network analyses were performed with MetaCore version 20.1 build 70000 (Clarivate Analytics, Philadelphia, PA), using differential gene expression outputs obtained from Seurat and SCENIC analyses.

CyTOF fcs files were normalized with fca_readfcs ^30^, (retrieved November 6, 2019). Following normalization, fcs files were processed further using Cytobank v.7.3.0, where gating was performed using DNA, event length, cisplatin, and CD45, to identify single live immune cells. Gated fcs files were exported and analyzed further using the cytofkit R package (v 1.12.0)^31^. Clustering was performed in cytofkit using the Rphenograph method with the CD45, CD19, CD1c, CD4, CD8a, CD16, CD123, CXCR3, CD14, CD127, CCR6, CD25, CD3, CD38, HLA-DR, and CD56 markers. CytofAsinh was selected as the data transformation method, t-distributed stochastic neighbor embedding (tSNE) was used as the dimensionality reduction method, k was set at 30, to analyze 120,000 cells (10,000 cells per sample; SA=8, HC=4).

All analyses were performed with R software version 3.5.1 unless indicated otherwise. Values are reported as means, standard deviation, percentages. Chi-square test was used in categorical data comparisons; t-test was used in continuous data, and the two-sample test for equality of proportions with continuity correction for proportions. All p-values were adjusted by FDR or Bonferroni methods unless indicated otherwise. P values <0.05 were considered significant.

## Results

### Subject Characteristics

Subjects with SA (n=5) and HC (n=3) had similar age and sex distribution (Table 1). Subjects with SA had higher BMI (p=0.04). All study subjects had minimal cigarette exposure history, SA (n=2), and HC (n=0), and the two SA former smokers had one pack-year history of cigarette smoking each; none were actively smoking. All subjects with SA were using a combination of inhaled corticosteroids and long-acting β_2_ -agonists. Additional therapies included montelukast, tiotropium, and omalizumab. Despite lower mean and median pulmonary function values in SA, there were no statistical differences compared with HC. All subjects with SA fulfilled the severe asthma definition by EPR-3 guidelines ^32^.

**Table 1.** Subject Demographics

|  | Severe Asthma (n=5) | Healthy Control (n=3) |
| --- | --- | --- |
| Age (years) | 43 ± 17 | 38 ± 15 |
| Sex (Female: Male) | 4:1 | 2:1 |
| **Race (n)** |  |  |
| Caucasian | 3 | 1 |
| African-American | 0 | 1 |
| Asian | - | 1 |
| Other | 2 | 0 |
| Hispanic (n) | 3 | 1 |
| BMI (Kg/m<sup>2</sup>) | 36 ± 6 | 23 ± 4 |
| Smoking (n) | 2 | 0 |
| Pack years | 1 each | - |
| **Therapies % (n)** |  |  |
| Inhaled corticosteroids | 100 (5) | - |
| Long-acting $\beta_2$-agonist | 100 (5) | - |
| Montelukast | 60 (3) | - |
| Tiotropium | 40 (2) | - |
| Omalizumab | 20 (1) | - |
| FEV<sub>1</sub> pre-bronchodilator (% Predicted) | 80 ± 18 | 91 ± 7 |
| FEV<sub>1</sub>/FVC | 78 ± 5 | 82 ± 14 |
| FeNO (ppb) | 23 ± 14 | 23 ± 16 |
| Absolute blood eosinophils (cells/microliter) | 260 ± 196 | 6 ± 11 |

Abbreviations: BMI=Body mass index; FEV<sub>1</sub>=Forced expiratory volume 1 second; FVC=Forced vital capacity; FeNO=Fractional exhaled nitric oxide.

### Single-cell RNAseq analysis of PBMCs

To investigate transcriptional changes at the single-cell level in unstimulated cells and in response to poly I:C stimulation, we isolated PBMCs from patients with SA and HC. Isolated PBMCs were exposed to poly I:C (stimulated) or plain culture media (unstimulated) for 24 hours before single-cell isolation using the DropSeq method ^19^. Similar numbers of single-cell transcriptome attached to microparticles (STAMPs) were retrieved per condition (unstimulated: 600 ± 0; poly I:C: 500 ± 295, p=0.34). The number of input reads ranged between 30.7 × 10^6^ to 84.8 × 10^6^. The number of uniquely mapped reads ranged between 42% and 79%, with no differences between groups of conditions (p=0.63 and p=0.32), respectively (Supplementary Table 2).

**Table 2.**
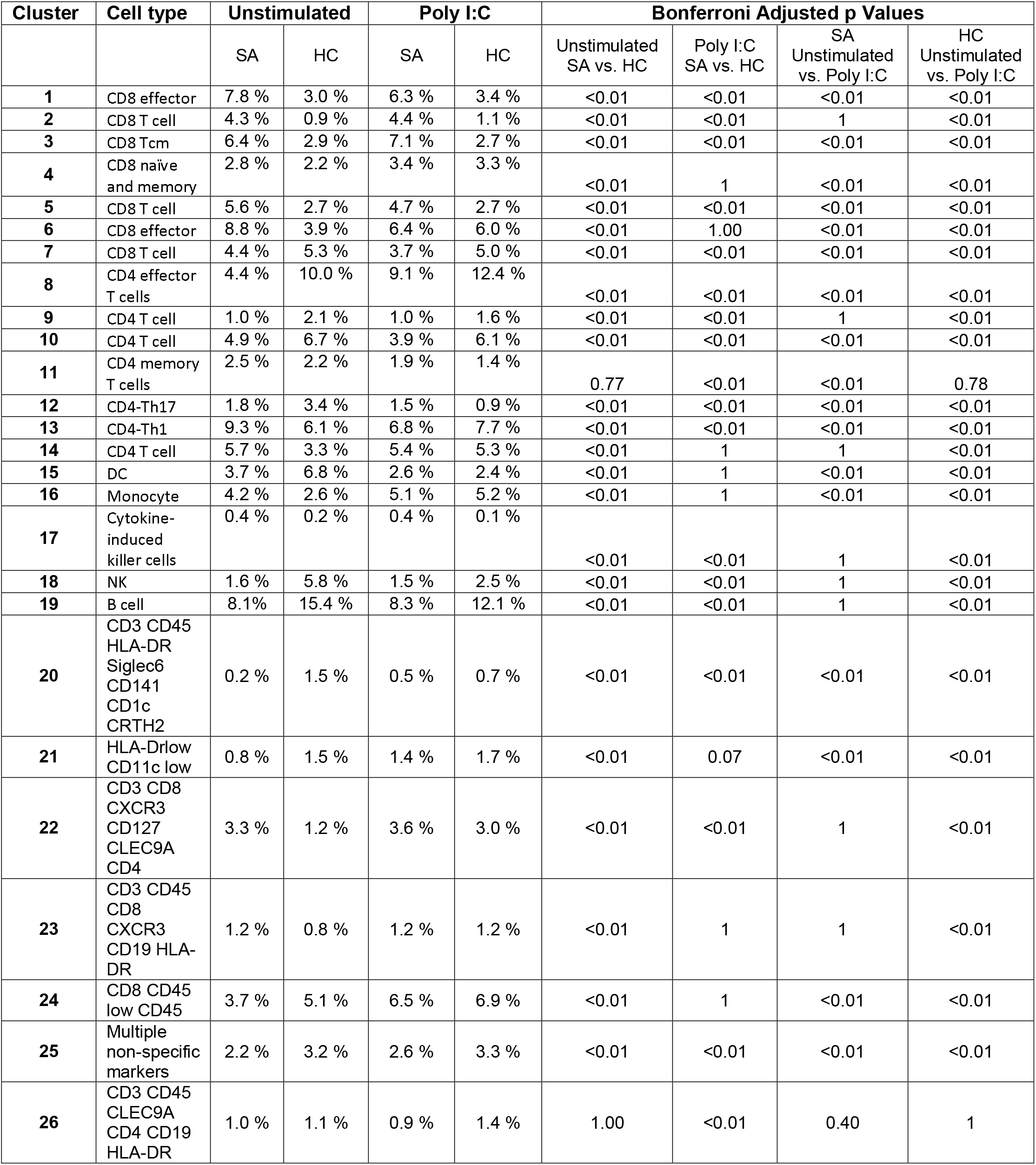
CyTOF cluster percentages by condition.

A mitochondrial gene expression threshold of 5% was used to select cells for downstream analysis, resulting in a total of 9,584 cells. Genes with high variability (n=2,000) were used before scaling gene expression data (Supplementary Table 3). A regression analysis using scaled data was performed using RNA counts, percentages of mitochondrial gene expression, and batch to remove unwanted variation sources. An independent analysis using cell cycle scoring analysis of scaled data did not show any particular bias toward a cell cycle-specific phase. Consequently, cell cycle variation was not included in the regression analysis. Elbow plot and the jackStraw procedures were used to identify principal components (PCs) and yielded similar results. Fifteen PCs were used to identify groups of correlated gene sets to define cell clusters (Figure 2A).

**Figure 2.**
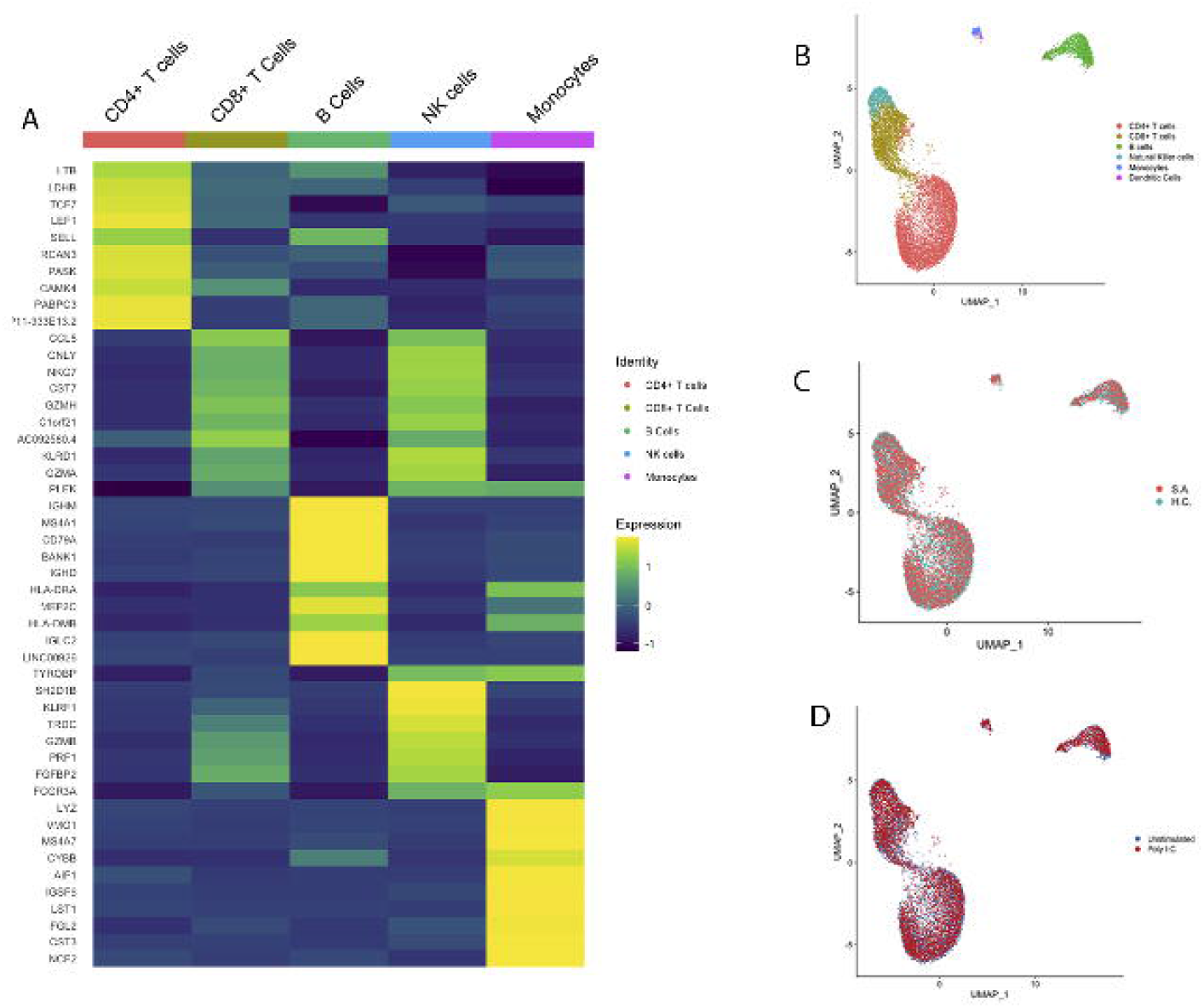
Single-cell RNAseq of PMBCs Identifies Distinct Clusters of Cells. A. Top ten cluster markers for the five main cell types in all cells (n=9,390). B. UMAP of all cell clusters including dendritic cells (n=9,414). C. UMAP of all cell clusters by disease status, severe asthma (SA) (n=(n=6,099) and healthy control (HC) (n=3,315). This figure demonstrates a similar distribution of cells across patients with severe asthma and healthy controls. D. UMAP of all cell clusters by stimulation status, unstimulated (n=4,283) and poly I:C (n=5,131). This figure demonstrates a similar distribution of unstimulated and poly I:C stimulated cells.

Specific cell markers were used to classify distinct groups of cells using singleR ^33^. Further manual curation complemented the singleR assignment. Total cell counts per condition were: 4,283 unstimulated and 5,131 poly I:C stimulated cells. The number of cells per disease status was: 6,099 for subjects with SA and 3,315 for HC. In both SA and HC subjects, CD4 positive T cells were the predominant group of cells, 54% in both, while dendritic cells (DCs) were the smallest group and ranged from 0.2% to 0.4%, respectively. Given the absence of enough DCs in SA subgroups (1 cell in unstimulated, 3 cells in poly I:C), we were unable to perform analyses across all conditions. Consequently, we concentrated our analyses in 9,390 cells to include CD4+ T cells, CD8+ T cells, NK cells, B cells, and monocytes.

Genes associated with specific cell identity in all cells for CD4+ T cells, CD8+ T cells, Natural Killer (NK), B cells, and monocytes are summarized in Supplementary Table 4. The heatmap in Figure 2A illustrates the top ten representative cell cluster biomarker genes. These biomarkers illustrate the cell-specific transcriptional programs in PBMCs. UMAPs of single-cell cluster distribution, cell clusters by disease, and stimulation status, Figures 2B-D, demonstrate even distribution of cell types by disease and stimulation status.

### Unstimulated Single Cells of Severe Asthmatics Display a Pro-inflammatory State

To identify gene-expression differences between SA and HC in each specific cell subset in unstimulated cells, we compared the PBMCs of subjects with SA and HC. Unstimulated cells in SA and HC had a similar distribution (Figure 3A). Transcriptional differences between unstimulated PBMCs are summarized in Supplementary Table 5. Among the most representative transcripts, the Janus kinase (*JAK1*) was highly expressed in CD4+ T cells, CD8+ T cells, NK cells, B cells and monocytes of SA compared to HC. CD4+ T, CD8+ T, NK and B cells of SA also showed high expression of the long non-coding RNA (lncRNA) *NEAT1*, associated with Th2 differentiation ^34^ (Figure 3B). In SA, the pro-inflammatory cytokine *IL32* was also highly expressed in all cell types except monocytes (Figure 3B).

**Figure 3.**
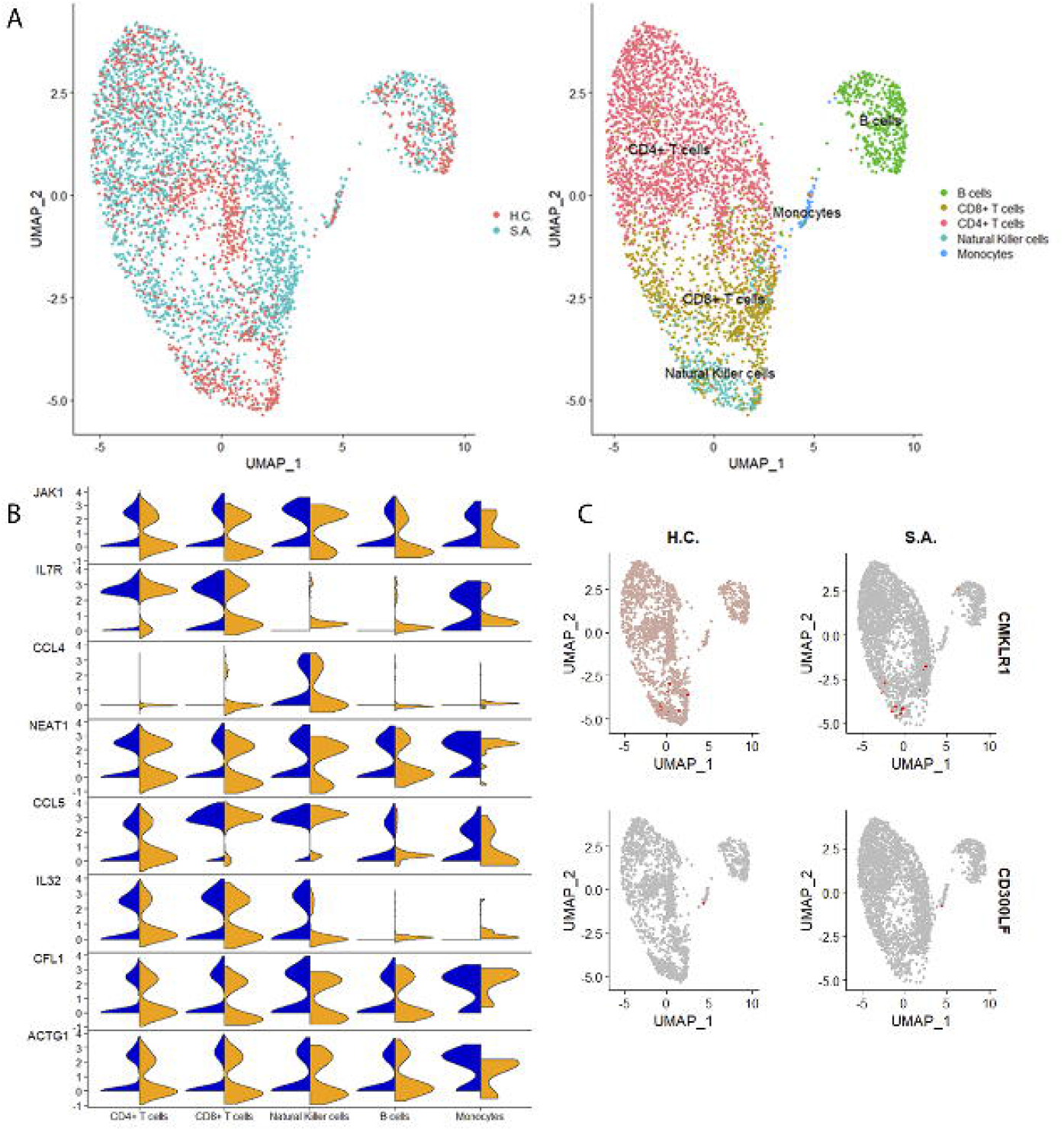
Single-cell RNAseq of unstimulated cells. A. UMAP of unstimulated PBMCs demonstrates a similar distribution of cells between severe asthma (SA) and healthy controls (HC). B. Several pro-inflammatory transcripts including *JAK1, IL7R, CCL4, NEAT1, CCL5, IL32, CFL1* and *ACTG1* are highly expressed in cells from patients with SA (blue) compared to HC (orange). C. Expression of the anti-inflammatory transcripts *CMKLR1* and *CD300LF* was lower in SA than HC.

CD4+ T and B cells, and monocytes of SA displayed higher expression of *CCL5*, a known eosinophil chemoattractant and activator ^35^. In comparison, CD8+ T cells of SA had a higher expression of *CCL4* (MIP-beta). In monocytes of SA, the chemerin chemokine-like receptor 1 (*CMKLR1*), the receptor for the chemoattractant adipokine chemerin/RARRES2, and the omega-3 fatty acid-derived molecule resolvin E1 ^36^, and *CD300LF*, a receptor involved in the negative regulation of mast cell activation ^37^, were both downregulated. In NK cells of SA (Fig. 3C), however, *CMKLR1* was upregulated, demonstrating cellular heterogeneity in this receptor’s expression.

To determine which pathways were enriched in unstimulated cells from SA, we performed enrichment analyses by cell type (Supplementary Table 6). This analysis demonstrated enrichment of pathways involved in cytoskeletal remodeling and cell adhesion in CD4+ T, CD8+ T, NK, and B cells from SA (FDR p <0.01). *CCR3* signaling in eosinophils was also enriched in CD4+ T, CD8+ T, NK, and B cells from SA; this pathway includes *CCL5, PFN1, CFL1, ACTB1, MYL12A, MYL12B* and *CALM1* (FDR p <0.01). A network of genes formed by *CD2, CD3, CD94, GRB2, CALM2, SET, ETS1, SRGN, JAK1*, and *H3FB*, involved in NK cell cytotoxicity was also highly expressed in SA (FDR p <0.01). NK cells from SA also demonstrated lower expression of multiple genes involved in phagocytosis and antigen presentation, including *IGKC, HLA-DRA*, and *HSP90AB1* (FDR p<0.01). Enrichment of B cells showed that B cells of SA were enriched for *CXCR4* signaling (FDR p <0.01). Thus, CD4+ T, CD8+ T, NK, and B cells of SA demonstrated similar pro-inflammatory changes.

In contrast with other cell types, monocytes of SA demonstrated enrichment for a Th17 pathway in asthma (FDR p<0.01), and the main gene ontology (GO) processes were regulation of immune system process and cellular response to cytokine stimulus (FDR p<0.01). Also, in contrast with CD4+ T, CD8+ T, NK, and B cells of SA, monocytes showed decreased expression of *CFL1* and *PFN1*, involved in the CCR3 signaling in eosinophils pathway. Across all unstimulated cell types of SA, monocytes demonstrated unique pathway enrichment patterns. Single genes and pathways with pro-inflammatory effects were highly expressed in patients with SA, as well as decreased expression of genes involved in resolution or negative regulation of inflammation. Together, these transcriptional changes are consistent with a pro-inflammatory status in unstimulated single cells of SA.

### PBMC Responses to Poly I:C Stimulation Demonstrate Induction of IFN Pathways in Severe Asthma

We hypothesized that PBMCs of SA might have decreased interferon-stimulated genes (ISGs) expression following exposure to poly I:C. To test this hypothesis, we measured genome-wide RNA expression in single-cells by DropSeq in poly I:C stimulated PBMCs from SA and HC. In contrast with our hypothesis, poly I:C stimulation was associated with higher expression of interferon-stimulated genes (ISGs) in SA (Supplementary Table 7). Several ISGs and genes involved in interferon pathways shared the same behavior across cell types in SA, except for *BIRC3, EDN1, FAM65B, IFITM2, ISG20*, and *SELL*, that were downregulated in B cells (Supplementary Table 7). Despite these B-cell specific differences, the overall response across cell types in SA was associated with higher expression of multiple interferon pathway genes than HC.

The plant homeodomain finger protein 11 (*PHF11*), a positive regulator of Th1 cytokine gene expression ^38^, was highly expressed in monocytes, CD4+ T, CD8+ T, and NK cells of SA, despite the concomitant increased expression of the lnc *NEAT1* associated with Th2 differentiation ^34^. *IL7R*, part of the heterodimeric receptor for the thymic stromal lymphopoietin (TSLP), and *JAK1* were highly expressed on CD4+ T, CD8+ T and B cells from SA. Together, these findings suggest that multiple cells from SA have a robust induction of genes involved in both Th1 and Th2 inflammation.

To identify pathways and networks enriched in single cells following poly I:C stimulation, we performed enrichment analyses in differentially expressed genes between SA and HC (Supplementary Table 8). In all cell types of SA, immune pathways of IFN-alpha/beta signaling via *JAK/STAT* and/or IFN-alpha/beta signaling via mitogen-activated protein kinases (MAPKs) were enriched in comparison with HC. CD4+ cells of SA also had increased expression of genes involved in the generation of memory CD4+ T cells, including *TRAC* and *TRBC2*, subunits of the T-cell antigen receptor, *IL2RG, IL7RA, JAK1, CCR7* and *SELL*. In CD8+ T and B cells of SA, a pathway involved in negative regulation of HIF1A function was enriched for genes that were highly expressed in SA and included *RACK1, HSP90AB1, HSP90B1, HSPA8, PSMA7, UBA52, SKP1*, and *CCNL1* (FDR p<0.01). Enrichment by disease of downregulated genes in CD8+ T cells of SA identified several genes involved in chronic obstructive pulmonary disease, including *CCR7, HCK, HSPA1L, IGHD, HCK*, and *TGFB3* (FDR p<0.01). Furthermore, in monocytes of SA, genes involved in antigen presentation by MHC class I, including *CLEC4A, HSP90B1, HSPA8, NPM1*, and *OLR1*, were downregulated in contrast with monocytes of HC (FDR p<0.01). Thus, single cells from SA demonstrated robust induction of IFN signaling pathways and induction of memory in CD4+ T cells, with downregulation of pathways involved in antigen presentation.

Given the decreased expression of antigen presentation genes in monocytes, in contrast with other cells, we focused on cell-specific changes in phagocytosis and antigen presentation pathways. We found that CD4+ T cells demonstrated enrichment of antigen presentation genes; however, elements of antigen presentation pathways and phagocytosis were differentially expressed across cell types and disease status, with *HLA-DRA, IGKC*, and *IGHM* demonstrating low expression on cells from SA. In contrast, other genes, including *CD74, HSP90AB1, PDIA3*, and *PSME1*, were highly expressed in SA (Figure 4B). Therefore, genes involved in antigen presentation and phagocytosis had distinct expression patterns in SA in response to poly I:C stimulation. Similarly, several genes involved in the unfolded protein response and ubiquitination, including *DNAJA1, UBXN4, HSPA8*, and *TMBIM6*, showed a heterogeneous expression pattern in cells of SA (Figure 4B). Together these findings identify cell-specific differences in antigen presentation, phagocytosis, unfolded protein response, and ubiquitination pathways in cells from SA.

**Figure 4.**
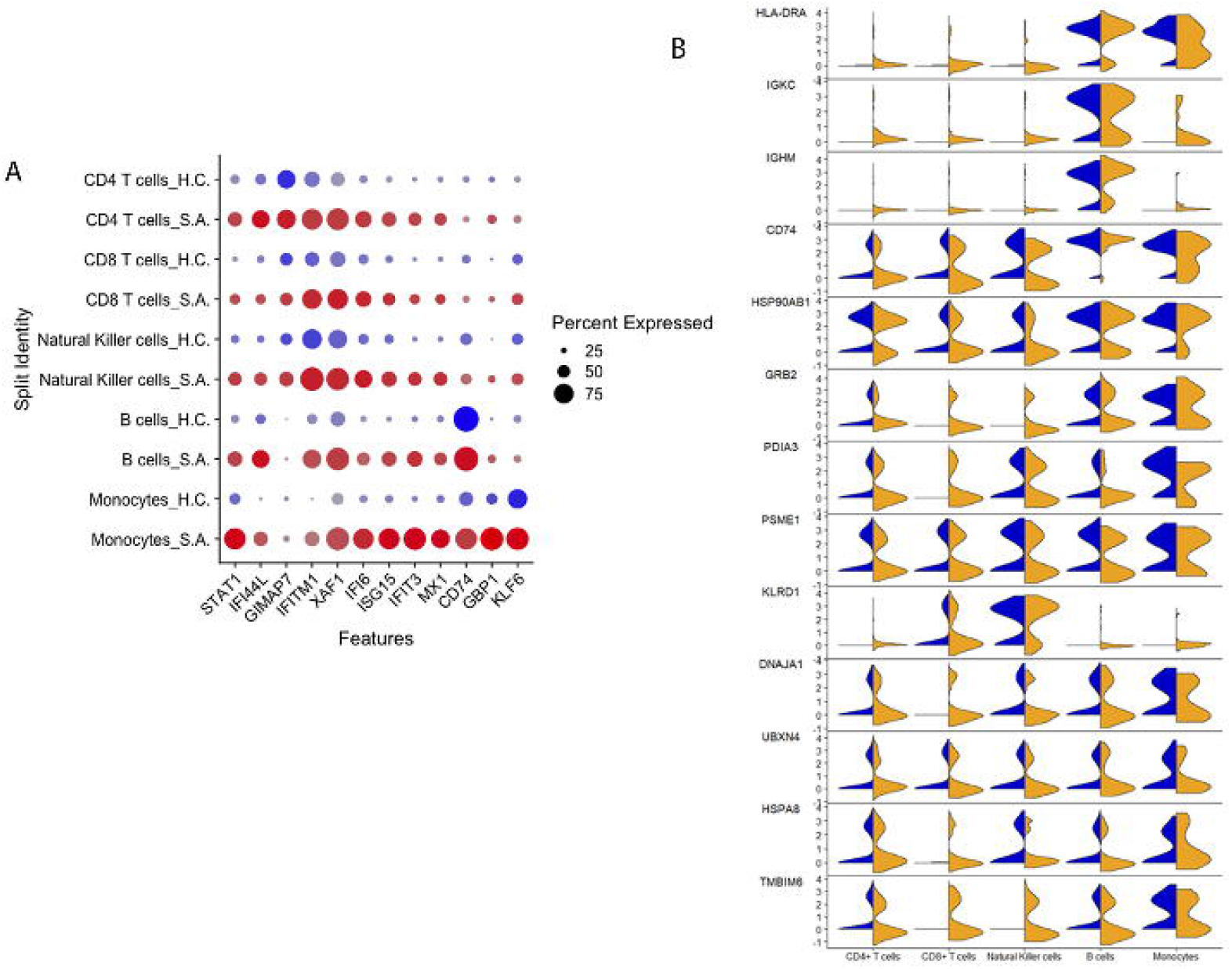
Single-cell RNAseq of PBMCs stimulated with poly I:C. A. Poly I:C stimulation let to a robust increase in the expression of STAT1 and multiple interferon stimulated genes in SA. B. Genes involved in antigen presentation including HLA-DRA, IGKC, IGHM, and CD74 had a heterogeneous expression across cell types and between SA and HC. Similarly, genes involved in ubiquitination and unfolded protein response also demonstrated a heterogeneous response across cells and disease status.

### Trajectory Analysis of CD4+ T Cells Identify Five Main Regulons in Response to Poly I:C Stimulation

To better understand IFN signaling pathways in response to poly I:C, we next examined the main transcription factors associated with the dynamic response to poly I:C stimulation in CD4+ T cells using pseudotime analysis with PHATE ^25^. PHATE embedding of CD4+ T cells was used to generate a pseudotemporal reconstruction of branching lineages ^26^ and identified the trajectory seen in Figure 5A. The trajectory inference was then correlated with CD4+ T single-cell transcripts to identify co-expressed genes and transcription factors correlated with pseudotime ^27^. This method identified 456 genes correlated with pseudotime at Bonferroni p<0.05 (Supplementary Table 9). Positively correlated genes were enriched for interferon type I signaling via JAK/STAT (FDR p<0.01) (Supplementary Table 10) consistent with the enrichment seen in CD4+ T cells of SA (Supplementary Table 8). In contrast, negatively correlated genes were enriched for the signal-recognition particle (SRP)-dependent protein targeting to the membrane during translation (FDR p<0.01) (Supplementary Table 10). Regulon analyses found a strong correlation between these genes and five transcription factors, *IRF1, STAT1, IRF7, STAT2*, and *IRF9* (Bonferroni p<0.05) (Figure 5C; supplementary Table 11). These transcriptional and master regulatory responses were more prominent in SA cells than in HC (Figures 5B and 5C). These observations suggest that SA is associated with robust induction of antiviral transcripts regulated by five main transcription factors, *IRF1, STAT1, IRF7, STAT2*, and *IRF9*.

**Figure 5.**
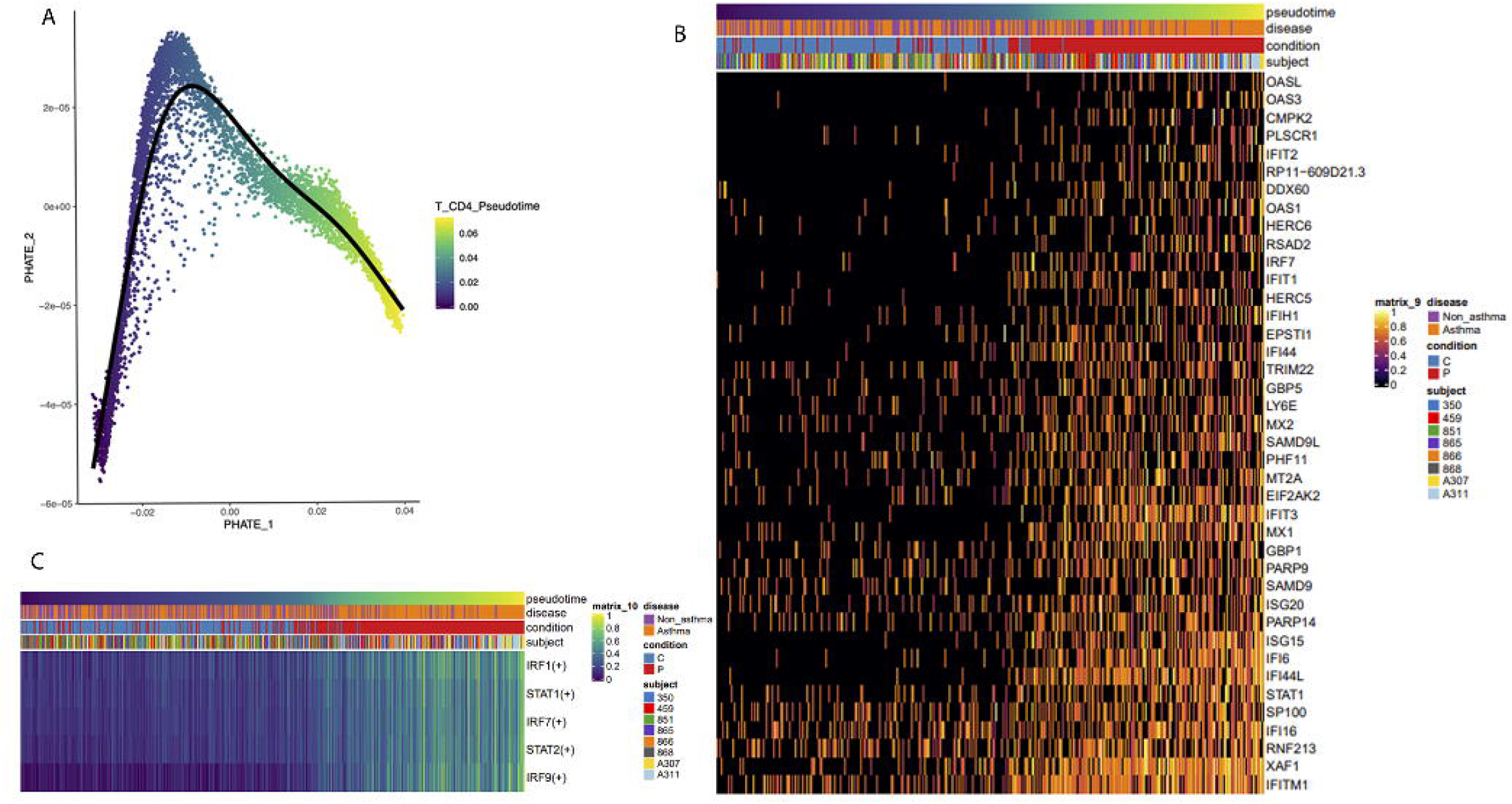
Pseudotime analysis of CD4+ T cells demonstrated a strong association with interferon signaling in severe asthma. A. PHATE analysis for pseudotemporal reconstruction of CD4+ T cells. B. Heatmap of transcripts correlated with pseudotime identified multiple interferon stimulated genes correlated with the response to poly I:C. C. Regulon analysis identified *IRF1, STAT1, IRF7, STAT2* and *IRF9* as the top five transcription factors positively correlated with pseudotime, and key regulators of positively correlated genes in response to poly I:C.

### CyTOF Analysis Identify Cell Abundance Differences in Severe Asthma

To identify and quantify PBMC subpopulations at baseline and following stimulation with poly I:C, we performed CyTOF staining of 21 immune cell surface markers (Supplementary Table 1). Clustering of 120,000 cells in 12 samples from the same subjects profiled with scRNA-seq (SA=4; HC=2) identified 26 cell clusters (Figure 6A and Table 2). Non-specific staining was seen in CRTH2 and CLEC9A; consequently, these markers were not used for clustering and did not affect cell type identification. Some clusters were formed by technical artifacts consistent with double staining (clusters 20, 22, 23, 25, and 26) and decreased staining (cluster 21). The presence of these clusters did not affect the identification of main cell subsets.

**Figure 6.**
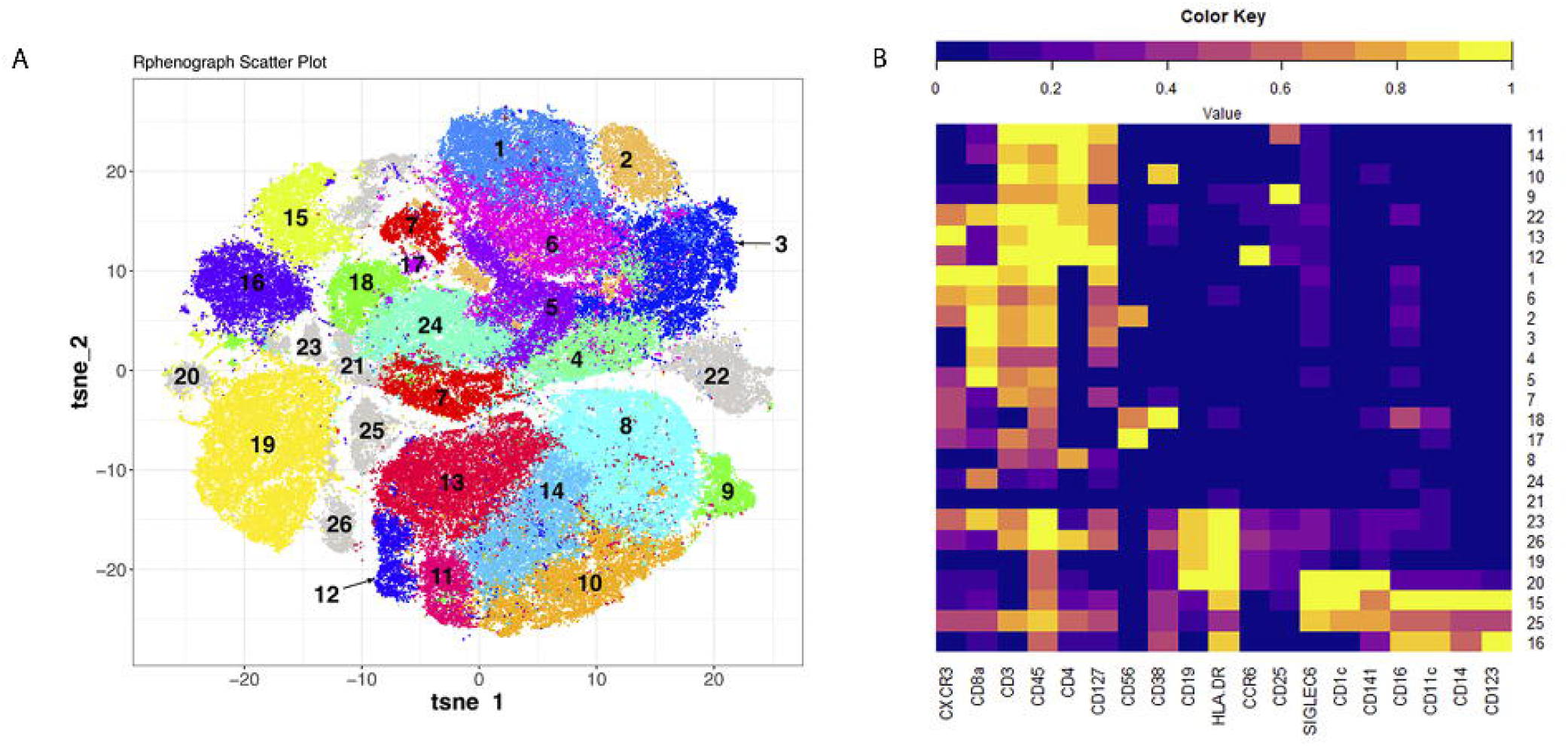
CyTOF analysis of PBMCs from the same subjects profiled with single-cell RNAseq identify a similar distribution of cells. A. tSNE plot of cell clusters on CyTOF. B. Heatmap of cell surface markers and clusters determined by CyTOF.

Consistent with scRNA-seq, CD4+ and CD8+ T cell clusters accounted for most of the cells, with an average of 70.8% in SA and 61.7% in HC. Compared to HC, unstimulated cells from SA had lower percentages of B cells (8.1% vs. 15.4%, p<0.01), NK cells (1.6% vs. 5.8%, p<0.01) and DCs (3.7% vs. 6.8%, p<0.01), while having a higher percentage of monocytes (4.2% vs. 2.6%, p<0.01). CyTof profiled a larger number of cells and enabled the identification of DCs, in contrast with the minimal number of DCs captured with DropSeq. Despite these differences in unstimulated cells, monocytes (5.1% vs. 5.2%, p=1) and DCs (2.6% vs. 2.4%, p=1) percentages were similar among SA and HC following poly I:C stimulation. Differences in cell counts between SA and HC persisted in B cells (8.3% vs. 12.1%, p<0.01) and NK cells (1.5% vs. 2.5%, p<0.01) following poly I:C stimulation.

CD8+ T cells were subdivided into seven clusters (1 to 7), and CD4+ T cells were also subdivided into seven clusters (8 to 14). Clusters of effector CD8+ T cells (1 and 6) and Th1 CD4+ T cells (13) were the most abundant in SA (Table 2). A decrease in effector CD8+ T cells was seen after poly I:C stimulation in SA, cluster 1 (7.8% vs. 6.3%, p<0.01), and cluster 6 (8.8% vs. 6.4%, p<0.01), while HC demonstrated an increase in both cluster 1 (3.0 vs. 3.4%, p<0.01) and cluster 6 (3.9% vs. 6.0%, p<0.01). Th1 CD4+ T cells in SA also decreased following poly I:C stimulation (9.3% vs. 6.8%, p<0.01) and increased in HC (6.1% vs. 7.7%, p<0.01). Together, these findings show several differences in relative cell abundance in SA compared to non-asthmatic controls. Specifically, higher CD8+ effector cells and Th1 CD4+ T cells in unstimulated conditions, was followed by a decrease of these two cell subsets after poly I:C stimulation. Thus, we observed quantitative differences in cell populations between SA and HC and significant changes between unstimulated and poly I:C stimulated cells.

## Discussion

In this study, we comprehensively analyzed the response of PBMCs to the TLR3 agonist poly I:C using scRNA-seq and CyTOF on samples from patients with severe asthma and HC. Contrary to our expectation, this study shows that cells of severe asthmatics do not have an impaired response to IFN stimulation. We found that stimulation of PBMCs from severe asthma with poly I:C led to a higher expression of ISGs than cells from HC. This observation suggests that the interferon pathways stimulated by poly I:C do not appear to be impaired in all patients with severe asthma. Furthermore, we identified quantitative differences in cell composition in severe asthma using CyTOF. These results provide a single-cell resolution map of PBMC function and abundance in patients with severe asthma.

We hypothesized that specific cells of severe asthmatics have an impaired response to IFN based on previous studies showing impaired antiviral immunity in asthma ^11,12,17^. The induction of ISGs across CD4+ T, CD8+ T, NK, B cells and monocytes of severe asthma was robust and supported by pathway enrichment analyses demonstrating multiple instances of immune responses linked to IFN, specifically, IFN-alpha/beta signaling via JAK/STAT and IFN-alpha/beta signaling via MAPKs.

Several reasons may account for these differences between our findings and previous studies. First, we explored a highly specific subset of patients with severe asthma. The response to poly I:C may vary across asthma endotypes, particularly in exacerbation-prone subgroups ^39–41^. Second, we examined PBMCs, while other studies have evaluated the airway compartment ^11,12^. Third, the stimulus used may also be responsible for differences in these responses. Therefore, our results must be interpreted in the context of an *in vitro* model rather than mimicking immune changes involved in asthma exacerbations.

A potential explanation for the enhanced response to poly I:C may be related to the observed increase in baseline expression of multiple pro-inflammatory and signaling genes, including *JAK1*, in severe asthma. We had previously identified a baseline increase in *STAT1* expression in patients with recurrent infectious exacerbations ^9^. JAK1 and STAT1 converge in an immune pathway involved in response to IFN and inflammation ^42^. Although the *JAK1-STAT1* pathway may explain differences in response to IFN seen here and in our previous study, the association between asthma and this pathway is complex, as it may be affected by inhaled corticosteroids and may represent a distinct asthma endotype ^43,44^. Thus, transcriptomic changes in the *JAK1-STAT1* pathway may account for some of the results seen in response to poly I:C stimulation.

We found that *STAT1*, in combination with *IRF1, IRF7, STAT2*, and *IRF9*, were the main regulators enriched in response to poly I:C in our study. Previous studies have shown that the transcriptional role of *STAT1* in asthma appears to be associated with corticosteroid insensitivity in asthma and Th1 bias ^43,44^. Similarly, *IRF1* has been associated with corticosteroid insensitivity ^45^, and *IRF9* associated with interferon networks in the airway epithelium ^*46*^. In contrast, *IRF7* has been associated with allergic airway inflammation through regulation of type 2 innate lymphoid cells ^47^. The observed increase in JAK1 expression in unstimulated cells, combined with a prominent role for *STAT1* in steroid and poly I:C responses, suggests that the *JAK1-STAT1* pathway holds clues to the complex interaction between asthma, therapeutic response to steroids, and antiviral responses.

In addition to finding increased expression of *JAK1* in unstimulated cells of severe asthmatics, we found high expression of pro-inflammatory genes including *IL32*, a cytokine with pro-inflammatory and antiviral activity ^48^; genes involved in Th2 inflammation, including *CCL5* and the lnc *NEAT1* ^34,35^; and *CCL4*, involved in asthma exacerbations ^49^. These changes are also accompanied by the downregulation of the *CMKLR1* receptor and *CD300LF* involved in anti-inflammatory mechanisms ^36,37^. Together, these single-cell transcriptomic changes indicate pro-inflammatory status at baseline in patients with severe asthma.

Although our central question pertaining to the response of PBMCs to poly I:C, a synthetic IFN trigger ^18^ and consequently focused on antiviral pathways, we found cell-specific heterogeneity in antigen presentation, phagocytosis, and unfolded protein response in cells of severe asthmatics. Antigen presentation in asthma has multiple implications in disease pathogenesis and intercellular communication ^50–52^, while the unfolded protein response is dysregulated in asthma and airway inflammation ^53,54^. Thus, cell-specific changes in antigen presentation and unfolded protein response pathways may underlie disease heterogeneity in asthma.

An additional insight from our studies that may underlie the disease heterogeneity of asthma is the quantitative difference in cell composition in severe asthma using CyTOF. We found significant differences in multiple cell population percentages between patients with severe asthma and HC using CyTOF. Although variation in eosinophil counts has led to the identification of eosinophilic asthma endotypes ^3,55,56^, cell abundance of other immune cell populations has not been explored in detail. However, it may be associated with distinct asthma features ^57^. The observation of changes in unstimulated and poly I:C stimulated cells, particularly in CD8+ T cells and Th1 cells, combined with the specific functional changes seen in sc-RNA-seq, may amplify the effect of the functional cellular changes in response to environmental stimuli when quantitative cell changes are combined with functional changes. This observation deserves further prospective evaluation to determine whether quantitative and qualitative changes are responsible for increased between-patient heterogeneity.

We are aware of the limitations of studying a highly selected group of individuals with asthma. However, we leveraged two single-cell methods to assemble a dataset with thousands of cells representing patients at the high end of the disease severity spectrum. Although these transcriptional changes need to be studied in milder asthma, our findings inform future study design, including the need to enrich dendritic cells to power single-cell analyses in the PBMC compartment. We sought to complement the use of single-cell RNAseq with CyTOF of cells from the same individuals to enable the interpretation of both functional and quantitative changes in response to poly I:C. These two complementary methods, based on mRNA and protein detection, create a precise profile of single cells in severe asthma. Although these findings may not extrapolate to all patients with asthma, we expect to see similar, albeit less dramatic, changes in patients with mild and moderate asthma.

Response to interferons is an essential component of immunity, and impairments in response to interferons have been associated with asthma pathogenesis ^9,11–17^. The use of two single-cell methods to analyze the response of PBMCs from severe asthmatics to poly I:C, a synthetic IFN trigger, enabled the identification of robust ISG changes and quantitative changes in CD8+ and Th1 cells. The presence of increased JAK1 expression at baseline in SA may hold clues to these responses. Together, these observations provide evidence of qualitative (scRNA-seq) and quantitative (CyTOF) differences in severe asthma that may be associated with disease pathogenesis. The implications of these findings are that single-cell heterogeneity across patients in severe asthma may be associated with disease heterogeneity. Future studies using a combination of quantitative and qualitative single-cell methods in asthma cohorts can help validate the role of interferon pathways in the immune heterogeneity of asthma.

## Data Availability

RNAseq data will be available through the NIH Sequence Read Archive (SRA)

## Acknowledgments

M.F.S. is supported by a Miguel Servet type I contract from Instituto de Salud Carlos III, Fondo de Investigación Sanitaria (Spain). Susan Ardito for administrative support.

## Abbreviations

SA: Severe asthma
PBMCs: Peripheral blood mononuclear cells
Poly I:C: Polyinosine–polycytidylic acid
dsRNA: Double-stranded RNA
IFNs: Interferons
scRNA-seq: Single-cell RNA sequencing
CyTOF: Mass cytometry
HC: Healthy controls
YCAAD: Yale Center for Asthma and Airway Disease
PCA: Principal component analysis
UMAP: Uniform manifold approximation and projection
GO: Gene ontology
STAMPs: Single-cell transcriptome attached to microparticles
NK: Natural killer
DC: Dendritic cell
ISG: Interferon stimulated genes

